# SARS-CoV-2 Fusion Peptide-Directed Antibodies Are Elicited by Natural Infection and Can Mediate Broad Sarbecovirus Neutralization

**DOI:** 10.1101/2025.03.01.25323010

**Authors:** Alex L. Roederer, Chia Jung Li, Eunice Lim, Yi Cao, David H. Canaday, Stefan Gravenstein, Alejandro B. Balazs

**Affiliations:** Ragon Institute of MGH, MIT, and Harvard, Cambridge, MA, 02139, USA; Case Western Reserve University School of Medicine, Cleveland, OH; Geriatric Research Education and Clinical Center, Louis Stokes Cleveland Department of Veterans Affairs Medical Center, Cleveland, Ohio; Center of Innovation in Long-Term Services and Supports, Veterans Administration Medical Center, Providence, Rhode Island; Division of Geriatrics and Palliative Medicine, Alpert Medical School of Brown University, Providence, Rhode Island, USA; Brown University School of Public Health Center for Gerontology and Healthcare Research, Providence, Rhode Island

**Keywords:** COVID-19, SARS-COV-2, broad neutralization, fusion peptide, stem helix, neutralizing antibodies

## Abstract

Studies have demonstrated that repeated mRNA vaccination enhances the breadth of neutralization against diverse SARS-CoV-2 variants. However, the development of antibodies capable of neutralizing across the Coronavirinae subfamily is poorly understood. In this study, we analyze serum samples to determine their neutralization breadth and potency and identify their antigenic targets. Using a cohort of older individuals and healthcare workers, we track correlates of broad neutralizing responses, including fusion peptide (FP) antibody elicitation. We find that although broadly neutralizing responses are often a result of RBD-specific antibodies, a rare subset of donors produce FP-specific broadly neutralizing responses. Interestingly, FP-specific antibodies are not observed in COVID-naive individuals irrespective of vaccination regimen, but rather, they occur following natural infection or vaccine breakthrough. This study highlights which epitope targets underpin broadly neutralizing antibody responses to coronaviruses and suggests that existing vaccines are insufficient to promote the elicitation of FP-directed broadly neutralizing coronavirus antibodies.

## Introduction

SARS-CoV-2 first appeared at the end of 2019 in Wuhan, China and quickly spread across the world, with an estimated 778 million documented infections leading to over 7 million documented deaths^1^, though this is likely to be a gross underestimate^2^. The global impact of this virus resulted in the development of multiple monoclonal neutralizing antibodies and vaccines to limit disease severity. The spike protein is the predominant target of neutralizing antibodies against SARS-CoV-2. The spike is composed of two subunits, the S1 which contains the receptor binding domain (RBD) and the N-Terminal Domain (NTD), and the S2, which contains the highly conserved Fusion Peptide (FP), and the fusion machinery which contains the conserved Stem Helix (SH). The RBD is the predominant target of neutralizing antibodies given its role in binding to the ACE2 receptor on host cells^3^. In the second year of the pandemic, numerous monoclonal antibodies targeting the RBD were approved for treatment but have since been rendered ineffective due to escape by SARS-CoV-2 variants^4–6^. This escape highlights the urgent need for preventive therapies including vaccines that will more effectively target future variants.

COVID-19 vaccination has been critical for prevention of severe disease from SARS-CoV-2 worldwide. The first generation of COVID vaccines^7–10^ were based on the Wuhan strain of SARS-CoV-2 spike protein, containing 2 proline-stabilizing mutations that were previously used to stabilize other related spike proteins^11^. Numerous SARS-CoV-2 variants have arisen over the last four years, which display immune evasion and changes in viral infectivity^12–18^. The mRNA vaccine has been reformulated multiple times, first to a bivalent composition of Wuhan and BA.5 variant spike sequences^19–22^, and then to a monovalent XBB.1.5 spike sequence^23–26^, and more recently to a monovalent KP.2 spike sequence^27^.

Despite these reformulations being highly effective against individual variants they were directed toward, the virus continued to mutate with the JN.1 and KP.2 variants escaping XBB.1.5 boosted sera effectively^18,28,29^. At their peak, JN.1 and offshoot variants including KP.2 made up over 96% of all sequences being submitted to the GISAID database^30^. A majority of mutations in variants have occurred in the RBD region with JN.1 and KP.2 having 28-29 of their 58-61 mutations in the RBD, driving the escape of the humoral immune response^28–32^.

Despite the plethora of vaccines, the virus has continued to evade humoral immune responses due to the immunodominance of RBD-directed antibodies and the selective pressure on this region. Notably, the FDA met in the summer of 2024 to formulate a vaccine for 2025^33^, suggesting that COVID vaccines will continue to be updated annually, much like those for Influenza, potentially costing billions each year^34,35^. Understanding the nature of rare broadly neutralizing responses to SARS-CoV-2 in patients is crucial for the development of next-generation vaccines that can uniformly elicit broadly neutralizing coronavirus antibodies.

Ideally, coronavirus broadly neutralizing antibodies would neutralize all existing SARS-CoV-2 variants and be cross-protective against strains likely to emerge in the future. Therefore, future vaccines could target highly conserved regions within the spike protein. There are several highly conserved regions within the RBD among SARS-CoV-2 variants that can be targeted by neutralizing antibodies^36,37^. However, it has been theorized that the mutational landscape in the RBD is incredibly vast^38^, and strong immune pressure has consistently selected for the RBD escape mutations found in existing variants. The SH is conserved among betacoronaviruses while the FP is conserved among all coronaviruses. Multiple groups have identified broadly cross-reactive and neutralizing antibodies that target these less common epitopes^39–46^. Given its conservation across even distantly related coronaviruses, the FP represents an ideal target antigen for vaccine design, however it is unclear how these neutralizing antibodies to FP are elicited.

To understand the development of broadly neutralizing antibody responses, we characterized donor sera with widely different neutralizing potency and breadth for binding to conserved antigenic sites within the spike protein. Interestingly, we find that the most potent neutralizers have the highest RBD, SH and FP titers despite similar spike IgG ELISA titers as less potent donors. Furthermore, among broad neutralizers we find that RBD and FP specific antibodies correlate with neutralizing titer to SARS-CoV-2. To further understand the make-up of a broadly neutralizing antibody response, we selectively depleted antibodies to the RBD, FP and SH. We find that while RBD antibodies are predominantly responsible for broad neutralization, as others have seen^41,47–49^, there exist rare donors with neutralizing FP antibodies that contribute significantly to breadth, however, FP antibody development is limited in most donors. Importantly, we find that FP antibodies develop following natural infection, and that additional vaccination leads to a decline in FP-specific antibody titers. Taken together, these results highlight the shortcomings of existing vaccination regimens to elicit highly cross-reactive, broadly neutralizing antibodies.

## RESULTS

### Broad neutralizers have extensive breadth across related Betacoronaviruses

To investigate the breadth of neutralization, a cohort of 382 donor samples collected at different timepoints after vaccination were evaluated for WT, BA.1 or BA.5 spike pseudovirus neutralization (**Figure S1A**). Notably, some samples exhibited potent neutralizing activity against all three viruses, exceeding the limit of detection of our assay. Samples were binned into distinct groups based on neutralizing titers across the three variants (**Figure 1A**). Samples with pNT_50_ > 2000 against WT and > 1500 against both BA.1 and BA.5 were classified as *Broad-Potent*. Samples with at least one but not all pNT_50_ values exceeding these thresholds were classified as *Narrow-Potent*. Samples with pNT_50_ < 300 against WT and < 100 against BA.1 and BA.5 were classified as *Weak*, and all remaining samples were classified as *Moderate*. To determine whether these broadly neutralizing responses could extend to other human infecting coronaviruses, a subset from each group of neutralizers were further tested against multiple related betacoronaviruses and two common cold alphacoronaviruses (**Figure 1B**). We developed high throughput neutralization assays for pseudoviruses representing these diverse viruses. Given that 229E and MERS use human Aminopeptidase (hANPEP)^50,51^ and dipeptidyl peptidase-4 (hDPP4)^52,53^ as cell receptors, stable cell lines expressing these cell-surface proteins were developed and confirmed to express their respective receptors by flow cytometry (**Figure S1B, C**). Each cell line was susceptible to their respective pseudoviruses and neutralization assays with previously described neutralizing antibodies resulted in IC_50_ values that were similar to those previously reported^43,45^ (**Figure S1D**). The most potent and broad neutralizers exhibited activity against SARS-CoV-2 variants, SARS-CoV, and WIV1, but this high level of potency did not extend to other alphacoronaviruses including NL63, 229E or the betacoronavirus MERS.

**Figure 1:**
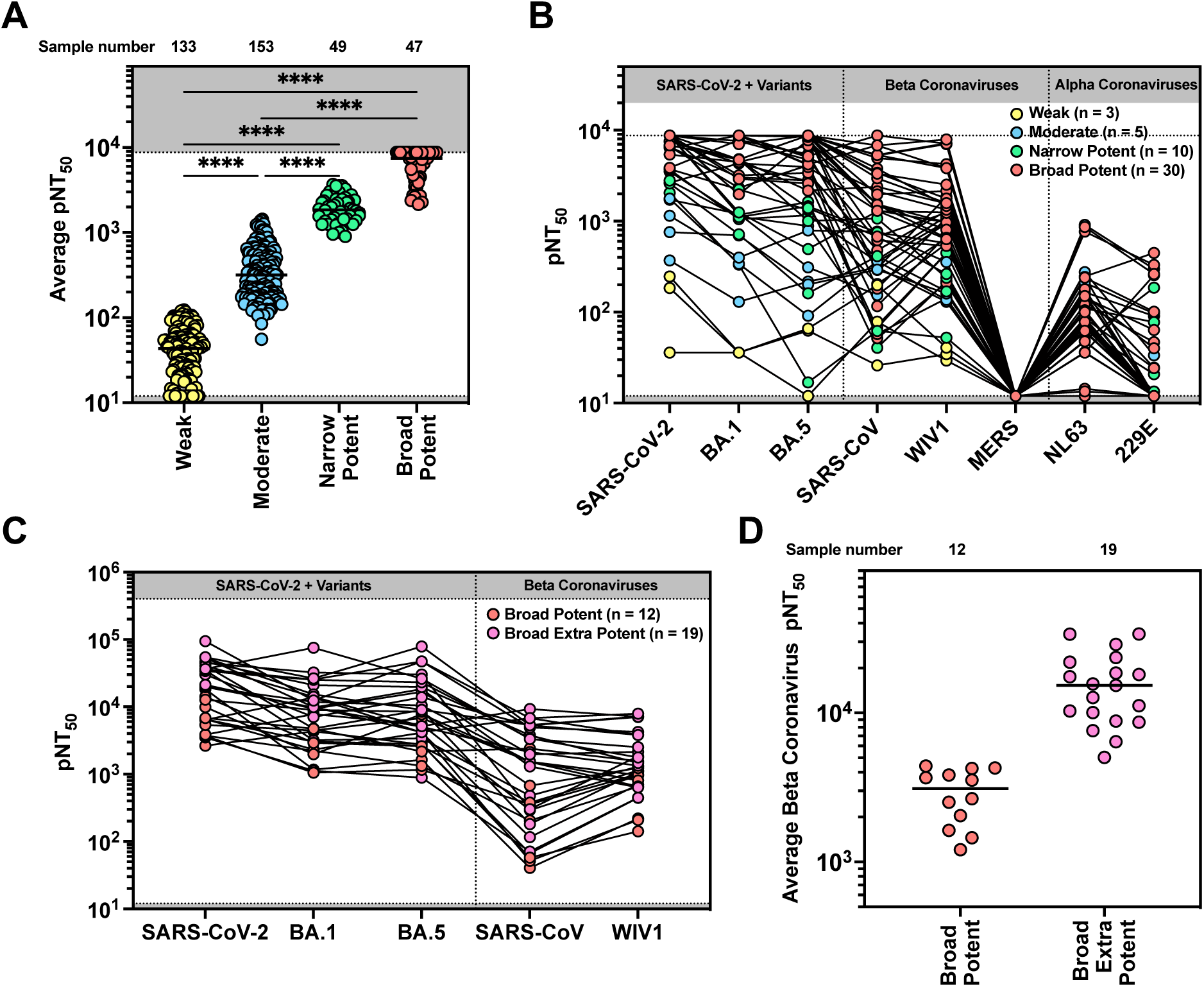
Broad neutralizers have extensive breadth across related Beta coronaviruses. (A) A cohort of 382 donor samples was divided into four groups based on neutralization potency (average of n = 2 technical replicates) against SARS-CoV-2 WT and the BA.1 and BA.5 variants, with a detection limit of pNT50 = 8748. Samples with pNT50 > 2000 against WT and > 1500 against both BA.1 and BA.5 were classified as Broad-Potent. Samples with at least one but not all pNT50 values exceeding these thresholds were classified as Narrow-Potent. Samples with pNT50 < 300 against WT and < 100 against BA.1 and BA.5 were classified as Weak, and all remaining samples were classified as Moderate. Because data did not pass normality tests (Shapiro-Wilk p < 0.05), non-parametric analyses were used. pNT50 values were compared using the Kruskal-Wallis test followed by Dunn’s multiple comparisons test (* = P<0.0332, ** = P<0.0021, ***= P<0.0002, ****= P<0.0001). (B) A subset of donor samples from each group were subject to our high throughput neutralization assay against SARS-CoV-2, variants, Beta Coronaviruses, and other distantly related coronaviruses for neutralizing activity (average of n = 2 technical replicates). (C) A subset of broad-potent samples was further tested against Beta Coronaviruses (excluding MERS) with a much higher threshold for neutralizing activity (average of n = 2 technical replicates). (D) Sera from (C) were subdivided into broad-extra-potent and broad-potent groups based on average neutralizing activity across the tested viruses using the cutoff pNT50 = 5000. Because the data were approximately normally distributed (Shapiro-Wilk p > 0.05), neutralization titers were compared between the groups using an unpaired two-tailed t-test with Welch’s correction for unequal variances.

*Broad-potent* neutralizers were re-tested at a higher dilution to allow for more accurate assessment of maximum neutralizing titers, which enabled further subdivision into *broad-extra-potent* and *broad-potent* based on an average of pNT50 values against SARS-CoV-2 WT and variants, SARS-CoV, and WIV1 (**Figure 1C**). While *broad-potent* sera exhibited extensive activity against SARS-CoV-2 WT, BA.1 and BA.5 variants, it lacked substantial neutralizing titer against related Betacornaviruses SARS-CoV and WIV1 as compared to *broad-extra-potent* sera. Average pNT_50_ values were re-calculated using values obtained from the extended dilution range assays. *Broad-extra-potent* neutralizers had a significantly higher average pNT_50_ of 16,191 while *broad-potent* had an average of 2,956 (**Figure 1D**). Given the importance of prior-exposure to SARS-CoV-2 in the development of humoral immunity, we determined COVID-expeirence of each sample based on both anti-N ELISA results as well as otherwise unexplained increases in SARS-CoV-2 spike-binding antibody titers. When comparing average pNT50 across the three viruses, we found that COVID-experienced individuals exhibited significantly increased titers as compared to those who were COVID-naive (**Figure S1E**). Interestingly, we found that while samples from COVID-naive donors also exhibited all four levels of potency, the proportion of potent neutralizers (*narrow-potent, broad-potent, and broad-extra-potent*) was higher in COVID-experienced donors (**Figure S1F, Table S1-3**).

### Broad neutralizers harbor the highest levels of stem helix and fusion peptide antibodies

Coronaviruses are highly diverse and classified into four genera, alphacoronaviruses, betacoronaviruses, deltacoronaviruses and gammacoronaviruses. All coronaviruses enter cells using their spike proteins, but the species tropism and cell receptor usage varies. Aligning the spike sequences across all four genera by conservation^54^ highlights that the RBD, the target for most neutralizing antibodies, are not conserved across coronaviruses (**Figure 2A**). The most conserved regions include the FP and the SH. Sera from the five groups of samples were assessed for their ability to bind to the full-length spike, RBD, FP, or SH by ELISA. Anti-spike (**Figure 2B**) and anti-RBD (**Figure 2C**) antibody endpoint titers were significantly higher in the *broad-extra-potent* and *broad-potent* groups than in the *moderate* or *weak* groups, as expected. Notably, the *broad-extra potent* group exhibited slightly lower spike-binding titers but higher RBD-specific titers than the broad-potent group. Importantly, only the *broad-extra-potent* group displayed significantly elevated antibody titers against FP (**Figure 2D**) and SH (**Figure 2E**) compared with the *moderate* group, suggesting that the increased potency and breadth of these samples may derive from antibodies targeting these conserved regions.. To determine the influence of prior COVID exposure on the development of antibodies against these epitopes, we compared the activity of COVID-naive or COVID-experienced samples across epitopes (**Figure S2**). COVID-experienced samples had higher endpoint titers against FP and SH but lower titers against RBD across most groups. Interestingly, in the *broad-extra-potent* group, COVID-experienced samples showed significantly higher FP titers than COVID-naive samples, while titers against full-length spike, RBD, and SH did not differ significantly (**Figure 2F**). These results suggest that the enhanced neutralizing activity in COVID-experienced broad-extra-potent samples may be driven by FP-directed antibodies.

**Figure 2:**
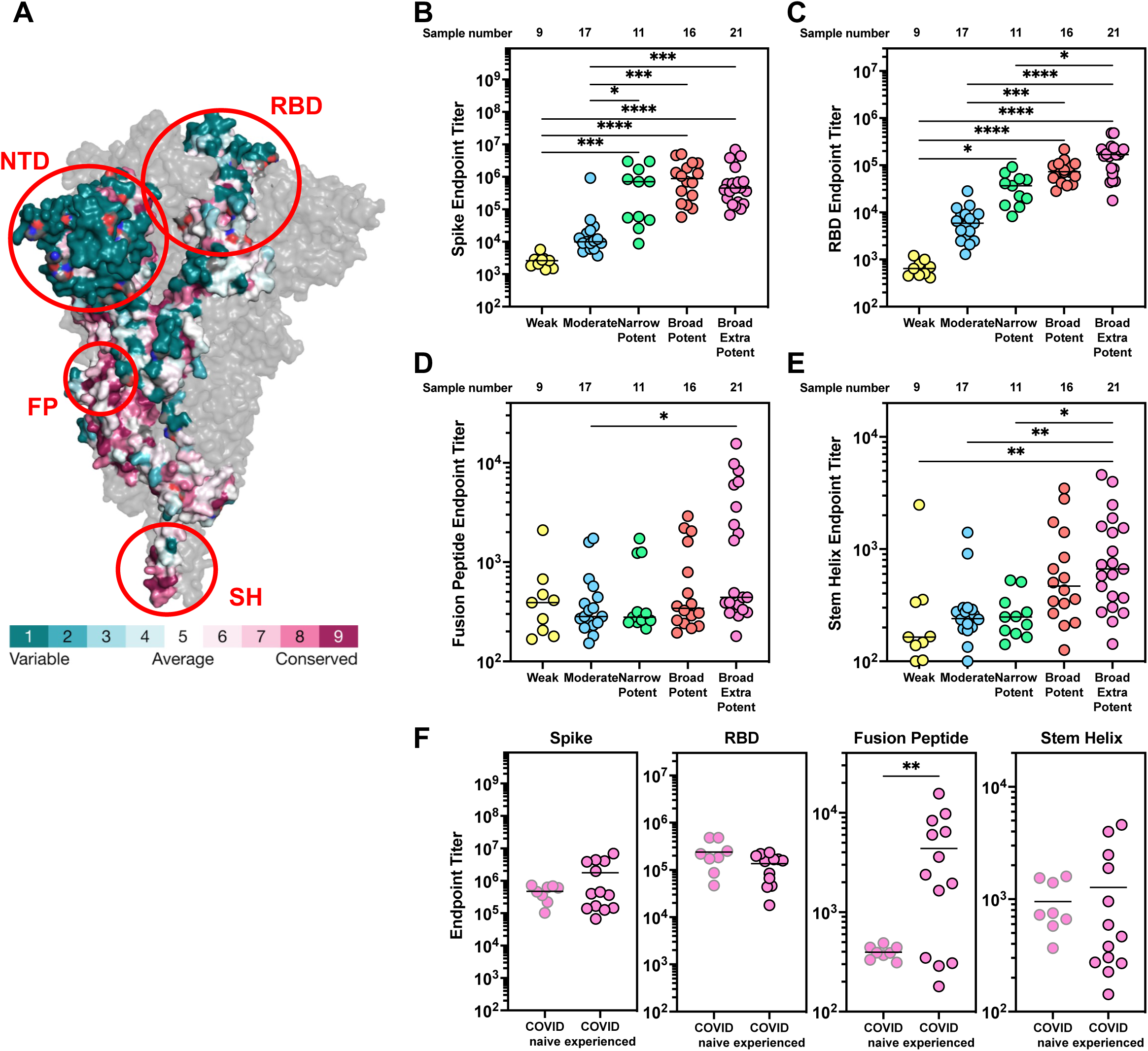
Broad-extra-potent sera have the highest levels of RBD, FP and SH antibodies. (A) Conservation of the amino acids within the spike protein generated using consurf^54^ from (PDB 6XR859 https://doi.org/10.2210/pdb6xr8/pdb). The sequences fed into the algorithm include 59 coronavirus spike sequences representing all four clades. RBD = Receptor Binding Domain, NTD = N-Terminal Domain, FP = Fusion Peptide, SH = Stem Helix. (B-E) Results from ELISA assessing the antibody endpoint titers of plasma from 21 broad-extra-potent, 16 broad-potent, 11 narrow-potent, 17 moderate and 9 weak donors. Antibody endpoint titers (average of n = 2 technical replicates) were assessed against an S2P Stabilized spike (B), Receptor Binding Domain (C), Fusion Peptide (D), or Stem Helix Peptide (E). Antibody endpoint titer represents the maximum interpolated dilution that can still bind the antigen. The endpoint titers were evaluated at a top dilution of 100-fold with seven additional 3-fold serial dilutions. The wells incubated with no plasma incubation were used as controls, and cut-off values (used for interpolation) were defined as 2.1X the OD450 of the controls. Representative data from two independent experiments is shown. Because data did not pass normality tests (Shapiro-Wilk p < 0.05), non-parametric analyses were used. Endpoint titer values were compared using the Kruskal-Wallis test followed by Dunn’s multiple comparisons test (* = P<0.0332, ** = P<0.0021, ***= P<0.0002, ****= P<0.0001). (F) Broad-extra-potent data from Figure 2B-D were further divided into two groups based on COVID history. Samples from COVID-naïve donors were outlined in gray, and those from COVID-experienced donors were outlined in black. Endpoint titers were compared between groups using an unpaired two-tailed t-test with Welch’s correction for unequal variances. ** = P<0.0021.

### RBD and fusion peptide specific antibodies correlate with neutralizing titer

To investigate the effect of antigen specific antibodies on neutralizing titer we selected 8 *broad-extra-potent* donors who had multiple serum collections over time (**Table S4**). Donors were older individuals with an average age of 71 as they came from a nursing home cohort. All donor samples were assessed for their antibody titers against spike, RBD, FP and SH to determine the amounts of antibody targeting each epitope. Known antibodies specific for each epitope were used to establish standard curves to interpolate Unit/mL values for each sample. A non-specific control antibody at the same serum IgG concentration was used to determine a cutoff for background binding to each antigen (**Figure 3, left**). All samples tested were above the cutoff for spike (**Figure 3A, left**) and RBD (**Figure 3B, left**), but only 11 samples reached the FP cutoff threshold and 9 samples reached the SH cutoff threshold (**Figure 3C, D**). To assess whether antigen-specific antibodies had an impact on SARS-CoV-2 neutralizing titer, correlation matrices were created for anti-spike (**Figure 3A**), anti-RBD (**Figure 3B**), anti-FP (**Figure 3C**) and anti-SH (**Figure 3D**) binding measurements. Correlation matrices only contained samples falling above their respective background levels to prevent bias by samples below the limit of detection. Despite the limited sample size, RBD and FP binding titers both correlated with the WT SARS-CoV-2 spike neutralizing titer, while spike and SH binding titers did not correlate with neutralization.

**Figure 3:**
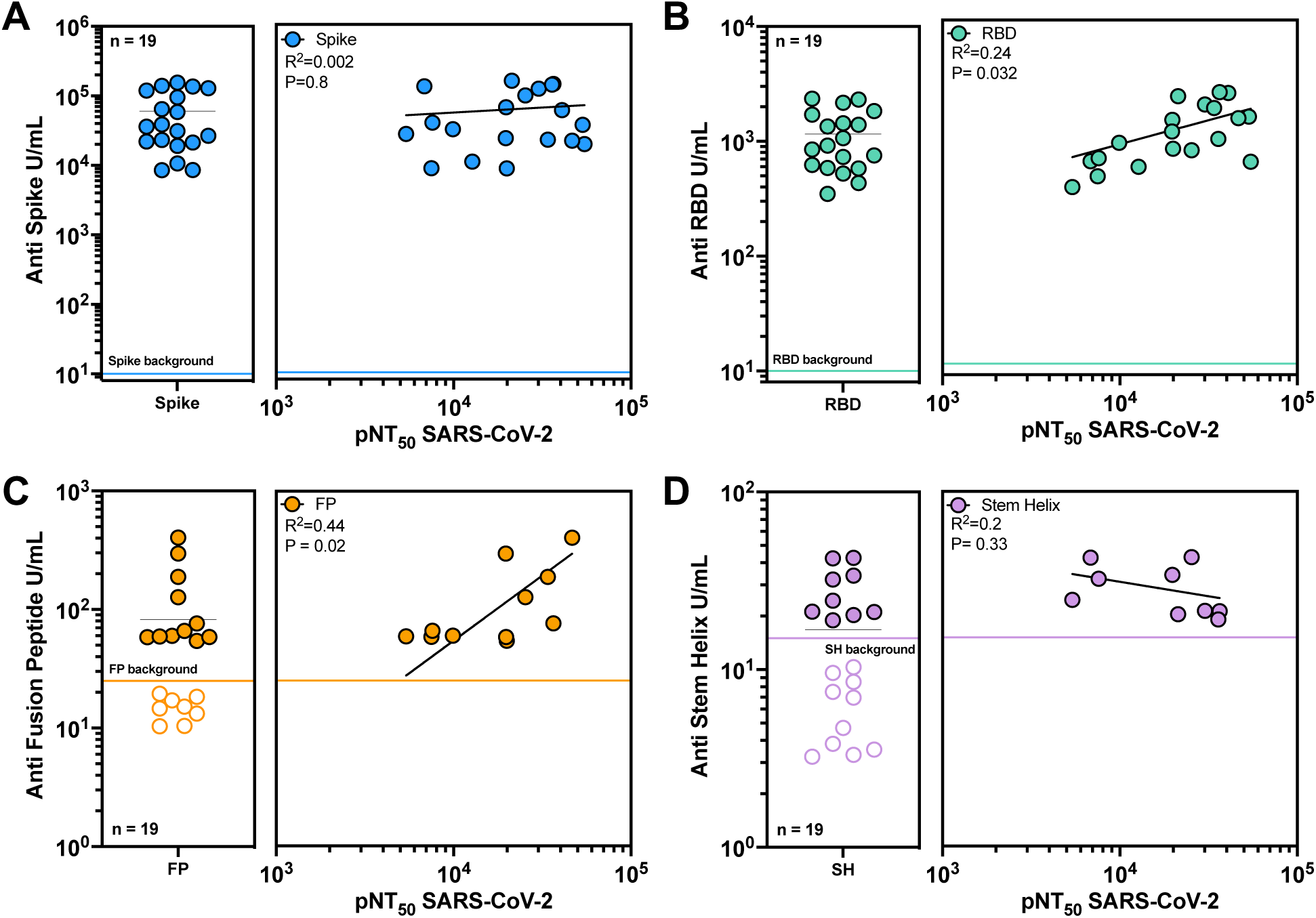
RBD and fusion peptide specific antibody titers correlate with neutralizing activity. (A-D, left) Unit/mL (U/mL) of antigen specific antibody was assessed for a cohort of broad and potent neutralizers (average of n = 2 technical replicates). All U/mL were calculated using an antigen specific ELISA and were defined as the equivalent reactivity to 1 µg/mL binding of the following antibodies: RBD182 for Spike (A) and RBD (B), 76E1 for FP (C), S2P6 for SH (D). A negative control antibody (2A10) at the same concentration as the serum dilution was included to determine cutoff values to define background serum binding. Data below the cutoff were excluded from subsequent correlation analyses. (A-D, right) Log-log regression analyses were performed on neutralization versus anti-Spike IgG (A), anti-RBD IgG (B), anti-FP IgG (C), and anti-SH IgG (D). Pearson correlations were performed and R2 and p values are indicated.

Given the correlations with neutralizing titer, we sought to understand the effect of vaccination on the development of antigen specific antibodies with our cohort. Generally, each vaccine dose increased SARS-CoV-2 spike and RBD specific antibodies, while antibody responses went down only after the first boost but remained high after subsequent doses. (**Figure S3A, B**). While antibodies targeting SH increased in some donors following vaccination, antibodies targeting FP did not change in this set of donors (**Figure S3C, D**). Unsurprisingly, SARS-CoV-2 vaccination showed no impact on the neutralizing titers of the alphacoronaviruses NL63 (**Figure S3E**) or 229E (**Figure S3F**), and no donor was able to neutralize MERS, irrespective of vaccination (**Figure S3G**). This suggests that vaccine-induced breadth extends to SARS-CoV-2 variants and closely related viruses, it does not extend to alphacoronaviruses or MERS.

### Fusion peptide antibodies can contribute to neutralization breadth

To investigate the contributions to breadth of RBD, FP, and SH-specific antibodies, we selectively depleted a subset of samples which had sufficient sera and high neutralizing activity in Figure 3, using SARS-CoV-2 RBD domain or peptides coupled to magnetic beads (**Figure 4A**). Mock depletions were performed by mixing the uncoupled beads with serum without antigen. Depletions were highly selective against the depleting antigen for both his-tagged WT SARS-CoV-2 RBD (**Figure S4A**), and maleimide coupled FP and SH (**Figure S4B**). We selected a subset of samples with the highest neutralizing activity, 8 of which potently bound FP and 6 of which potently bound SH, for depletion with each respective antigen (**Figure S4C, D**). Overall, depletion of FP or SH-specific antibodies had a minimal effect on the neutralizing titers of samples against both SARS-CoV-2 (**Figure S4E**) and SARS-CoV (**Figure S4F**). However, a few samples (FP3, FP6) had slightly reduced neutralizing activity against both SARS-CoV and SARS-CoV-2 when depleted for FP-specific antibody binding while a single sample had slightly reduced neutralizing activity against both viruses after SH Depletion (SH4).

**Figure 4:**
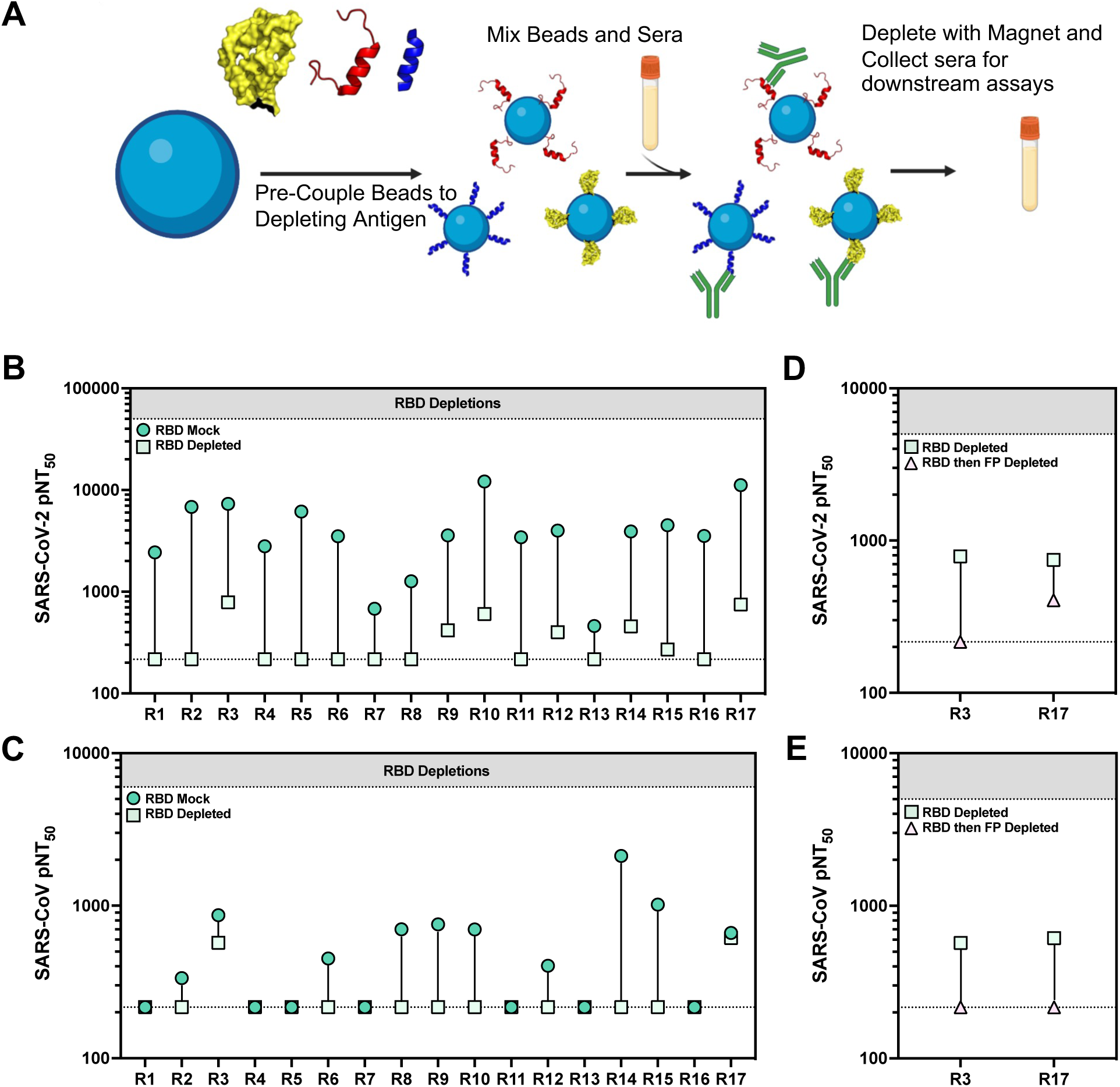
Fusion peptide antibodies contribute to neutralization breadth. (A) Schematic detailing our antigen depletion protocol. Briefly, his-tag or maleimide binding magnetic beads are pre-coupled to the antigen/peptide and mixed with serum. After an incubation the magnetic beads are pulled away using a magnet and serum is collected for downstream assays. Mock depletion is done by mixing serum and beads with no antigen (Created in BioRender). (B-C) Serum samples with high neutralizing activity were subject to RBD based depletion using his-tagged beads or mock depletion using beads with no RBD bound. Mock and depleted serum was tested for neutralizing activity (average of n = 2 technical replicates) against SARS-CoV-2 (C) and SARS-CoV (D). (D-E) Serum samples with remaining neutralizing activity against both SARS-CoV-2 and SARS-CoV from (B-C) were subject to further depletion with the fusion peptide and tested for remaining neutralizing activity (average of n = 2 technical replicates) to SARS-CoV-2 (D) and SARS-CoV (E) after double depletion.

It has been previously shown that SARS-CoV-2 RBD depletion of sera can remove the majority of neutralizing activity^47–49,55,56^. To determine whether this would also be the case with potent neutralizing sera, we depleted 17 potently neutralizing samples using SARS-CoV-2 RBD (**Figure S4G**). Of note, RBD-depleted serum samples 3,9,10,12,14,15, and 17 retained some neutralizing activity against SARS-CoV-2 (**Figure 4B**), while RBD-depleted samples 3 and 17 retained substantial neutralizing activity against SARS-CoV (**Figure 4C**). These results suggest that these samples harbored non-RBD, cross-reactive, neutralizing antibodies. As both sample 3 and 17 exhibited potent FP binding, the SARS-CoV-2 RBD-depleted samples were further depleted to remove FP-binding antibodies (**Figure S4H**). While sample 17 retained some neutralizing activity to SARS-CoV-2 (**Figure 4D**), both samples lost all neutralizing activity against SARS-CoV (**Figure 4E**), suggesting that FP-specific responses in these samples contributed to broad neutralizing activity.

### Fusion peptide specific antibodies arise only from natural infection

Given the variability of FP-specific antibody levels we measured across donor samples, we sought to determine when FP-specific antibodies arose. Using a cohort of pre-vaccine COVID-experienced serum samples (**Table S5**), we found that more severe COVID infections led to higher FP-specific antibody levels. (**Figure 5A**). However, we found no significant differences in FP titers regardless of age (**Figure S5A**), race (**Figure S5B**) or biological sex (**Figure S5C**).

**Figure 5:**
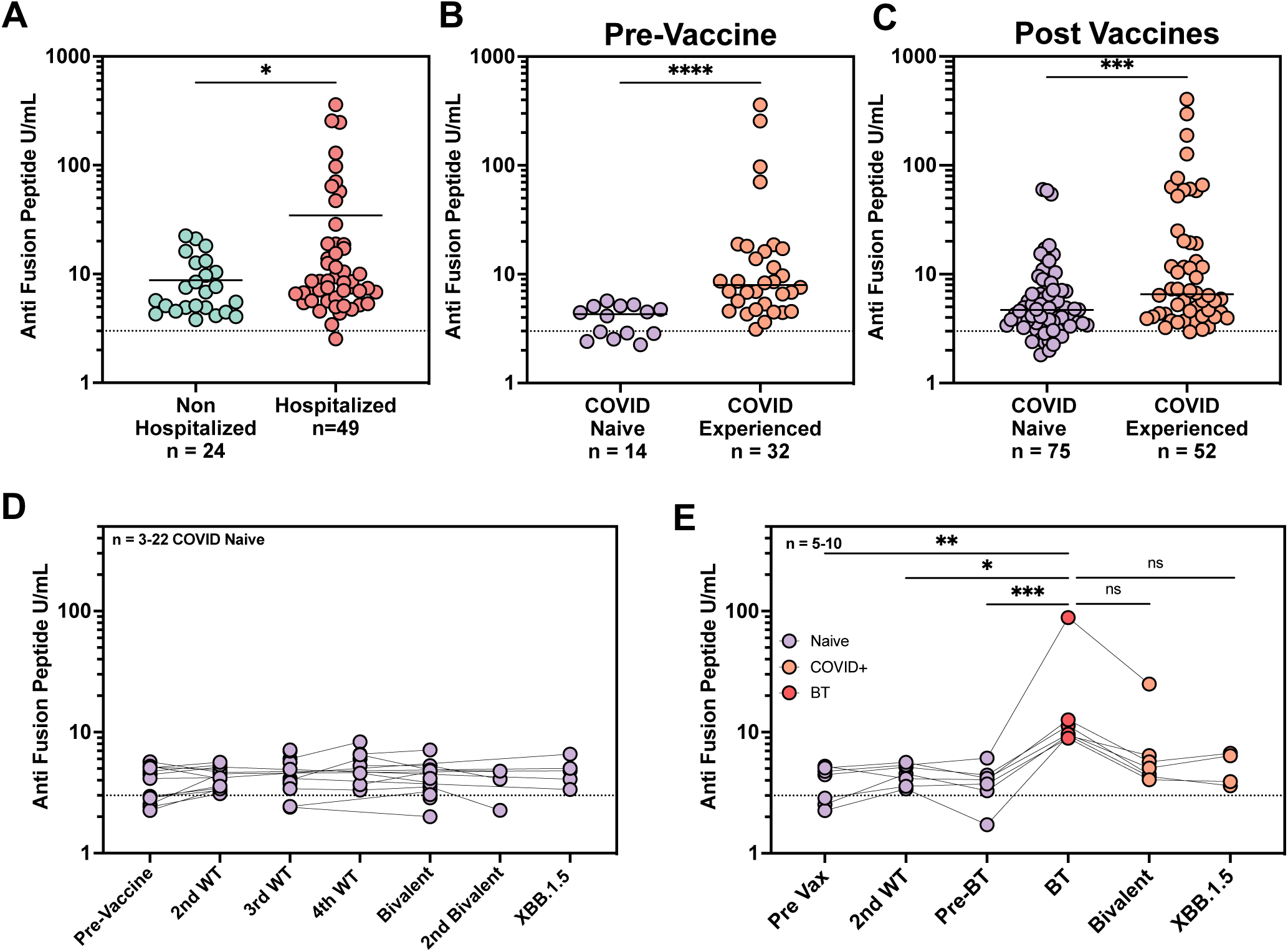
Fusion peptide antibodies develop in response to natural infection. (A) Anti-Fusion Peptide ELISA for a cohort of COVID-experienced pre-vaccination timepoints split into non-hospitalized and hospitalized (average of n = 2 technical replicates). Limit of detection line is defined as the background binding of a negative control antibody to the FP for that experiment. Because the data did not follow a normal distribution (Shapiro-Wilk p < 0.001), differences between these groups were analyzed using the Mann-Whitney U test (two-tailed). * = P<0.0332. (B) Anti-Fusion Peptide ELISA (average of n = 2 technical replicates) for pre-vaccination serum samples for donors who were naïve or who had been infected with COVID. A Mann Whitney Two tailed T-test was performed to assess significance. ****= P<0.0001. (C) Anti-Fusion Peptide ELISA (average of n = 2 technical replicates) for post-vaccination serum samples for donors who were naïve after 2-6 vaccines or who were COVID-experienced at the timepoint and had received between 2-6 vaccines. A Mann Whitney Two tailed T-test was performed to assess significance. ***= P<0.0002. (D) Fusion peptide ELISA (average of n = 2 technical replicates) was performed for a cohort of COVID naïve donors at various stages of vaccination. No significance was seen between groups in the development of FP specific antibodies. Limit of detection line is defined as the background binding of a negative control antibody to the FP for that experiment. (E) Anti-Fusion Peptide ELISA (average of n = 2 technical replicates) for longitudinal timepoints of donors who had a breakthrough infection between 2nd vaccine and bivalent. Pre-Breakthrough (Pre-BT) indicates the time point closest to breakthrough infection (BT) timepoint. A Kruskal Wallis one-way ANOVA was performed with Dunn’s correction for multiple comparisons (* = P<0.0332, ** = P<0.0021, ***= P<0.0002, ****= P<0.0001). For A-E U/mL is defined as the reactivity seen by 1 µg/mL of 76E1 antibody. Cutoffs were determined by binding of a negative control antibody (2A10).

We next sought to determine the effect of vaccinations on FP-specific antibody elicitation. Donor samples were gathered from aged-matched cohorts containing older individuals to test for FP-specific responses by ELISA (**Table S5**). COVID-naive and COVID-experienced donors were first compared to determine whether FP-specific antibodies could be detected in sera obtained before SARS-CoV-2 vaccines were available (**Figure 5B**). As expected, donors who were COVID-experienced had significantly higher levels of FP-specific antibodies than naive donors, suggesting that FP-specific antibodies were not common prior to infection. To assess FP-specific antibody responses in donors who had been vaccinated, samples from a vaccinated cohort of COVID-naive or COVID-experienced donors were analyzed by ELISA. Vaccinated COVID-experienced donors had significantly higher levels of FP-specific antibodies than those who were COVID-naive (**Figure 5C**). Separately, samples obtained from a longitudinal cohort of COVID-naive donors from pre-vaccination to XBB.1.5 booster (**Table S5**) were tested for FP-specific antibodies. Interestingly, all COVID-naive donors in this cohort had low levels of FP-specific antibodies irrespective of vaccination or boosting (**Figure 5D**). However, longitudinal samples obtained from COVID-naive donors who experienced breakthrough infection between their WT booster and bivalent vaccines, showed a significant increase in FP-specific antibodies after infection (**Figure 5E**). This cohort also exhibited an apparent decrease in FP-specific antibodies after bivalent vaccines and XBB.1.5 booster, though these decreases did not reach statistical significance. Together, these results suggest that COVID infection, rather than vaccination, plays an important role in the development of FP-specific antibodies. These findings suggest that mRNA vaccines do not effectively elicit FP-specific responses and may reduce these responses when they arise as a consequence of breakthrough infection.

## Discussion

We sought to uncover how broad neutralization of SARS-CoV-2 and related coronaviruses developed following vaccination or infection and assessed the contribution of various spike epitopes to broad recognition. To this end, we determined the neutralization potency of hundreds of donors to find those with the most potent and broad neutralization. We found that neutralization activity in sera from *broad-extra-potent* neutralizers predominantly arises from RBD-specific antibodies as has been previously reported^57^. However, *broad-extra-potent* neutralizers also had the highest levels of FP and SH-specific antibodies. Moreover, among the *broad-extra-potent* samples, only RBD and FP-specific antibodies correlated with neutralizing titer. Despite increases in spike and RBD antibodies that were observed over multiple vaccine administrations, there was no correlation between vaccination and the development of FP and SH-specific responses. In line with this, neutralization breadth driven by vaccination extended to SARS-CoV-2 variants and SARS-like viruses, but did not extend to more distantly related beta and alpha coronaviruses.

Depletion of SARS-CoV-2 RBD-specific antibodies drastically reduced neutralization potency of sera, while FP-specific depletion resulted in only minimal effects, suggesting that the vast majority of neutralization can be attributed to RBD-specific humoral immunity. However, rare donors maintained substantial neutralizing titers against SARS-CoV-2 and SARS-CoV after RBD-specific depletion, indicating the presence of cross-reactive neutralizing antibodies targeting an epitope outside of RBD. Serial depletion of RBD followed by FP resulted in loss of all neutralizing activity, confirming the potential for FP-specific antibodies to contribute to neutralization activity. Our results agree with previously published work showing that depletion of convalescent donor serum with a linear peptide including the FP reduced neutralization titers by only 20%^58^. Additionally, our results suggest the activity of RBD-specific antibodies can mask FP-directed neutralizing antibodies, and that RBD depletion is necessary to enable their detection. This is especially true given the wide range of neutralization potencies of monoclonal FP-specific antibodies against SARS-CoV-2^43–46^.

Given their potential as broad-spectrum treatments against coronaviruses, we sought to determine the source of FP-specific antibody development. Comparing COVID-naive and COVID-experienced cohorts, we found a clear, significant increase in FP-specific antibodies in COVID-experienced donors, suggesting that these antibodies primarily arise from natural infection. It has been shown that FP-specific antibodies lack binding to recombinantly expressed spikes, which contain either 2 or 6 proline mutations to stabilize the trimer^46^. Importantly, mRNA vaccines all encode spike proteins with the 2 proline mutations, which may contribute to their lack of elicitation of FP-specific antibodies in vaccinated, COVID-naive individuals. Indeed, we found that repeated mRNA vaccination did not promote FP-specific antibody development, whereas breakthrough infection significantly increased FP-specific responses. Interestingly, subsequent vaccination after infection in the breakthrough cohort largely reduced FP-specific antibody levels. While age, gender, and demographics were not correlated with FP-specific antibody development, greater COVID severity tended to induce higher levels of FP-specific antibodies. This may be due to the extremely high levels of spike-specific antibodies seen in severe COVID cases as we and others have shown a correlation between the amount of spike-specific antibodies and disease severity^59,60,60^.

Our study predominantly utilized samples from a cohort of nursing home donors who have been longitudinally profiled, highlighting a unique cohort which generally struggles to maintain effective immunity against continued SARS-CoV-2 evolution. However, we were able to find donors among these with very broad and potent neutralizing activity, demonstrating the potential to create effective humoral responses. These results also highlight the deficiencies of current mRNA vaccines for the generation of FP-specific broadly neutralizing coronavirus antibodies, suggesting that new vaccine designs may be needed for broader coverage. Updated domain-specific vaccines have demonstrated promising results in clinical trials^61^, however, these lack FP sequences. In contrast, S2-specific vaccines also have shown potential for protection and contain the FP-region which may improve targeting of this epitope^62,63^. FP-specific vaccines have been suggested for HIV as a means of specifically eliciting antibodies against this target^64,65^. Whether similar vaccine designs presenting coronavirus FP could elicit comparable humoral responses remains to be determined.

Although the COVID-19 pandemic has been declared over by the WHO^66,67^, there are still tens of thousands of cases and hundreds of deaths each day and the virus continues to evolve to evade humoral immunity^68^. Additionally, the ever-present threat of a new zoonotic transmission event, given the vast reservoir of SARS-like viruses that exist in animal populations, highlights the need to develop more broadly effective vaccines. Development of approaches capable of efficiently eliciting FP-specific antibodies would ensure the availability of effective prophylaxis against future pandemic coronaviruses.

### Limitations of the Study

This study is limited to the evaluation of serum neutralizing antibodies to the spike protein using a pseudovirus-based neutralization assay, and does not take into account neutralizing antibodies to other proteins including the N and M proteins which may contribute to neutralizing breadth. Additionally, spike ELISAs used 2P-stabilized recombinant proteins, which could underestimate the total spike antibodies. It is known that FP-specific antibodies bind 2P-stabilized spikes with lower affinity than the native conformation of spike^69^. Our experiments also utilized linear peptides to represent the FP and SH and therefore could be underestimating the binding to these epitopes in the context of the native conformation of spike, however, others^39,43–45^ have used similar approaches for monoclonal antibody binding and for sorting of individual memory B cells. Furthermore, we did not assess the role of vaccine-elicited cellular immune responses mediated by T cells and NK cells, which play a key role in limiting disease severity for vaccine recipients. We also did not assess other antibody-mediated functions such as complement deposition, antibody-dependent cellular cytotoxicity, or antibody-dependent cellular phagocytosis, which may contribute to protection even in the absence of neutralizing antibodies.

## RESOURCE AVAILABILITY

### Lead Contact

Further information and requests for resources and reagents should be directed to and will be fulfilled by Alejandro Balazs.

### Materials Availability

Plasmids generated in this study will be available through Addgene. Recombinant proteins, peptides, and antibodies are available from their respective sources.

### Data and Code Availability

This study did not generate sequence data. Data generated in the current study (including neutralization measurements and flow cytometric files) have not been deposited in a public repository but are available from the corresponding author upon request. The code used to make the phylogenetic trees can be made available from the corresponding author upon request.

## Data Availability

All data produced in the present study are available upon reasonable request to the authors

## ACKNOWLEDGEMENTS, FUNDING SUPPORT

We wish to thank Michael Farzan, PhD, for providing ACE2-expressing 293T cells. This work was supported by the Peter and Ann Lambertus Family Foundation. A.B.B. was supported by NIAID R01s AI174875, AI174276, AI129709-03S1 the NIDA Avenir New Innovator Award DP2DA040254, the NIDA Avant-Garde Award 1DP1DA060607, a Massachusetts Consortium on Pathogenesis Readiness (MassCPR) grant, CDC subcontract 200-2016-91773-T.O.2 and a grant from Coalition for Epidemic Preparedness Innovations (CEPI).

## AUTHOR CONTRIBUTIONS

**Conceptualization:** A.R., A.B.B.

**Methodology:** A.R., Y.C.

**Samples:** D.H.C., S.G., M.P

**Investigation:** A.R., Y.C., C.J.L, E.L, T.D.

**Data Curation:** A.R.

**Writing – Original Draft:** A.R.

**Writing – Reviewing and Editing:** A.R., C.J.L., A.B.B. edited the paper with input from co-authors.

## DECLARATIONS OF INTEREST

A.B.B. is a founder of Cure Systems LLC. S. G. and D. H. C. are recipients of investigator-initiated grants to their universities from Pfizer to study pneumococcal vaccines, Sanofi Pasteur and Seqirus to study influenza vaccines, Moderna to study respiratory infection, and GSK to study herpes zoster. S. G. is the recipient of an investigator-initiated grant to the university from Genentech on influenza antivirals; reports consulting for GSK, Janssen/Johnson&Johnson, Merck, Novavax, Moderna, Seqirus, Sanofi, and Vaxart; he has received honoraria for speaking for AstraZeneca, Pfizer, Seqirus, Sanofi; reports personal fees from Pfizer; and reports data and safety monitoring board fees from Longevoron and SciClone.

### Disclaimer

The contents do not represent the views of the U.S. Department of Veterans Affairs or of the United States Government.

## STAR METHODS

### EXPERIMENTAL MODEL AND SUBJECT DETAILS

#### Human subjects

Use of human samples was approved by Partners Institutional Review Board (protocol 2020P002274). The current analysis is part of an ongoing study in which NH residents are consented directly or through their legally authorized representative if needed and serially sampled before and after each SARS-CoV-2 vaccine dose. This analysis includes data collected between September 29, 2021, and June 7, 2023. Residents who received SARS-CoV-2 mRNA vaccines [(BNT162b2 (Pfizer-BioNTech) or mRNA-1273 (Moderna)] were included, and those who received the Ad26.COV2.S (Janssen) vaccine were excluded. Participants received their first booster dose 8–9 months after the primary vaccination series, their second booster 4 to 6 months after their first booster and third booster 4 to 7 months after their second booster. We report results from blood samples obtained at time points prior to and following one, two or three booster doses. All donor genders were self-reported.

#### Cell lines

HEK 293T cells (ATCC) were cultured in DMEM (Corning) containing 10% fetal bovine serum (VWR), and penicillin/streptomycin (Corning) at 37°C/5% CO_2_. 293T-ACE2 cells were a gift from Michael Farzan (Scripps Florida) and Nir Hacohen (Broad Institute) and were cultured under the same conditions. Confirmation of ACE2 expression in 293T-ACE2 cells was done via flow cytometry. hDPP4-293T and hANPEP-293T were produced in-house and expression of hDPP4 and hANPEP was confirmed via flow cytometry.

### METHOD DETAILS

To create variant spike expression plasmids, we performed multiple PCR fragment amplifications utilizing oligonucleotides containing each desired mutation (Integrated DNA Technology) and utilized overlapping fragment assembly to generate the full complement of mutations for each strain. PCR reactions were done with Platinum SuperFI II (Thermofisher) according to the manufacturer’s protocol for reaction size and thermocycler protocol. Importantly, we generate these mutations in the context of our previously described (Garcia-Beltran *et al.* 2020^56^) codon-optimized SARS-CoV-2 spike expression plasmid harboring a deletion of the C-terminal 18 amino acids that was previously demonstrated to result in higher pseudovirus titers. Assembled fragments were inserted into NotI/XbaI digested pTwist-CMV-BetaGlobin-WPRE-Neo vector utilizing the In-Fusion HD Cloning Kit (Takara) according to the manufacturer’s protocol. In-Fusion reactions were transformed into Mix and Go DH5Alpha cells (Zymo Research) according to the manufacturer’s protocol and colonies were grown overnight at 37°C to generate plasmid DNA purified using a standard miniprep protocol (Qiagen). All resulting plasmid DNA utilized in the study was verified by whole-plasmid deep sequencing (Illumina or Primordium Labs) to confirm the presence of only the intended mutations and maxi-prepped (Macherey-Nagel) to ensure high quality, high concentrated DNA.

#### Construction of fusion and stem helix peptides

All peptides used in this manuscript were synthesized by GenScript with a C terminal cysteine for maleimide conjugation. The following sequences were used for the Fusion Peptide and Stem Helix: SKPSKRSFIEDLLNKVTLADAGC (FP), and DSFKEELDKYFKNHTSC (SH). Peptides were resuspended in the manufacturer recommended buffer, aliquoted into tubes, and stored at -20°C when not in use.

#### SARS-CoV-2 pseudovirus neutralization assay

To compare the neutralizing activity of vaccine sera against coronaviruses, we produced lentiviral particles pseudotyped with different spike proteins as previously described (Garcia-Beltran *et al.* 2020^59^). Briefly, pseudoviruses were produced in 293T cells by PEI transfection of a lentiviral backbone encoding CMV-Luciferase-IRES-ZsGreen as well as lentiviral helper plasmids and each spike variant expression plasmid. Following collection and filtering, production was quantified by titering via flow cytometry on 293T-ACE2 cells. Neutralization assays and readout were performed on a Fluent Automated Workstation (Tecan) liquid handler using 384-well plates (Grenier). Three-fold serial dilutions ranging from 1:12 to 1:8,748 were performed for each serum sample before adding 125–250 infectious units of pseudovirus for 1 hr. Subsequently, 293T-ACE2 cells containing polybrene at 8 µg/mL were added to each well and incubated at 37°C/5% CO_2_ for 48-60 hrs. Following transduction, cells were lysed using a luciferin-containing buffer (Siebring-van Olst *et al.* 2013^70^) and shaken for 5 min prior to quantitation of luciferase expression within 1 hr of buffer addition using a Spectramax L luminometer (Molecular Devices) or a Pherastar with Microplate Stacker (BMG Labtech). Percent neutralization was determined by subtracting background luminescence measured in cell control wells (cells only) from sample wells and dividing by virus control wells (virus and cells only). Data was analyzed using Graphpad Prism and pNT_50_ values were calculated by taking the inverse of the 50% inhibitory concentration value for all samples with a neutralization value of 80% or higher at the highest concentration of serum.

#### Titering

To determine the infectious units of pseudotyped lentiviral vectors, we plated 400,000 293T-ACE2 cells per well of a 12-well plate. 24 h later, three ten-fold serial dilutions of neat pseudovirus supernatant were made in 100 μL, which was then used to replace 100 μL of media on the plated cells. Cells were incubated for 48 h at 37°C/5% CO_2_ to allow for expression of the ZsGreen reporter gene and harvested with Trypsin-EDTA (Corning). Cells were resuspended in PBS supplemented with 2% FBS (PBS+), and analyzed on a Stratedigm S1300Exi Flow Cytometer to determine the percentage of ZsGreen-expressing cells. Infectious units were calculated by determining the percentage of infected cells in wells exhibiting linear decreases in transduction and multiplying by the average number of cells per well determined at the initiation of the assay. At low MOI, each transduced ZsGreen cell was assumed to represent a single infectious unit.

#### hANPEP and hDPP4 293T Cell Line Production

To create cell lines that could be used in the high throughput neutralization assay we first cloned the known cellular receptors hDPP4 (MERS) and hANPEP (229E) into a lentiviral vector backbone from cDNA. Lentivirus was then produced using the same plasmids as the pseudovirus without the spike or fLuc-ZsGreen vectors. 293T cells were seeded at 400,000 cells/well one day before, media was removed and an equal quantity of lentivirus was added to cells. After 48 hours, cells were counted and diluted to 1 cell/well of 2 X 96 well plates. Single cell clones were then grown up for 2 weeks and assessed for surface expression of the indicated receptor by flow cytometry using the following stains CD26-PE-Cy7 (Biolegend 302713), or CD13-BV785 (Biolegend 301725). The highest expressing single clone was then grown up to make the cell line and frozen down in 90% FBS 10% DMSO for further uses.

#### Endpoint ELISA

Endpoint ELISAs were determined as follows: 96-well NUNC MaxiSorp ELISA plates (ThermoFisher) were coated with either 0.8 µg/mL purified S2P stabilized spike protein, 1 µg/mL purified RBD (kindly provided by Aaron Schmidt, Jared Feldman, and Blake Hauser) or 2 µg/mL synthesized Stem-Helix or Fusion-Peptide (GenScript). Plates were washed with a wash buffer consisting of 50 mM Tris (pH 8.0) (Sigma), 140 mM NaCl (Sigma), and 0.05% Tween-20 (Sigma). Plates were incubated with a blocking buffer consisting of 1% BSA (Seracare), 50 mM Tris (pH 8.0), and 140 mM NaCl for 30 min at room temperature, and then washed. Serum samples were diluted 1:100 with a dilution buffer consisting of 1% BSA, 50 mM Tris (pH 8.0), 140 mM NaCl, and 0.05% Tween-20. Samples were added to corresponding wells and incubated for 1 hr at room temperature, followed by washing. Anti-human IgG-HRP (1:25,000) specific antibodies (Bethyl) were then used to detect bound serum antibodies and were diluted as indicated. These were added to each plate and incubated for 30 min at room temperature. After washing, detection was done with a TMB 2-Component Microwell Peroxidase Substrate Kit (VWR). Substrate B and Sample were mixed at a 1:1 ratio following manufacturer instructions and then added to each well and incubated for ∼5 min before stopping with 3 M H2SO4. Buffer compositions, reagent concentrations and incubation times and temperatures were optimized in separate experiments for each analyte to maximize signal-to-noise ratio. Optical density (O.D.) was measured at 450 nm with subtraction of the O.D. at 570 nm as a reference wavelength on a SpectraMax ABS microplate reader. Antibody endpoint titer represents the maximum interpolated dilution that can still bind the antigen. The endpoint titers were evaluated at a top dilution of 100-fold with seven additional 3-fold serial dilutions. The wells incubated with no plasma incubation were used as controls, and cut-off values (used for interpolation) were defined as 2.1X the OD450 of the controls.

#### Quantitative ELISA

ELISAs were performed identically to endpoint ELISAs. A standard curve was generated using the following antibodies and the WT SARS-CoV-2 spike ectodomain (SinoBiologic) was used for coating at 0.8 µg/mL: RBD182 antibody as a control for Spike and SARS-CoV-2 RBD binding, S2P6 as a control for Stem Helix binding, and 76E1 as a control for Fusion Peptide binding. Anti-RBD and anti-spike antibody levels were calculated by interpolating onto the standard curve and correcting for sample dilution; one unit per mL (U/mL) was defined as the equivalent reactivity seen by 1 μg/mL of a SARS-CoV-2 RBD antibody (RBD182) which was isolated from human donor PBMCs). Anti-SH and anti-FP antibodies were calculated similarly; one U/mL of SH antibody was defined as the equivalent reactivity seen by 1 µg/mL S2P6 while one U/mL of FP antibody was defined as the equivalent reactivity seen by 1 µg/mL 76E1.

#### Depletion Protocol RBD

Dynabeads (Thermofisher) were washed three times according to the manufacturer protocol and resuspended in PBS + 0.05% BSA. 5 µL of serum was added to molar equivalents of purified RBD (kindly provided by Aaron Schmidt, Jared Feldman, and Blake Hauser). Mock depleted sera was added to an equivalent volume of PBS + 0.05% BSA. Serum and RBD or serum and PBS + 0.05% BSA were incubated at room temperature for 1 hour. Beads were added to a magnet for 5 minutes, the supernatant was removed, and beads were resuspended in the RBD + Serum or Mock depletion mixture before incubating for 30 minutes at room temperature. Depletions were then placed on a magnetic strip for 5 minutes and supernatant was collected for downstream assays. RBD depletion was confirmed by an ELISA of mock and depleted samples for binding to the RBD.

#### Depletion Protocol Stem Helix and Fusion Peptide

Maleimide activated beads (Cube Biotech) were washed three times according to the manufacturer’s protocols in coupling buffer. 3 mg of peptide was then added to the coupling buffer to a final volume of 1.5 mL in an Eppendorf tube. Beads were then resuspended in the coupling buffer with a peptide and incubated on a thermoshaker overnight at 4°C or at 20°C for 2 hours. Beads were then incubated on a magnetic strip for 2 minutes before being washed 3 more times in a coupling buffer and then once in 1 mL distilled water. Finally, conjugated beads were resuspended in 500 µL of storage buffer (Dissolve 2.1g of NaHCO3 in 150 mL water, adjust pH to 7.5 then add 1 mL sodium azide (5% w/v) and fill up to 250 mL with water) and stored at 4°C for future use. Before depletion, beads were washed 3 times with PBS + 0.05% BSA and then resuspended in 5 µL Serum diluted in 45 µL PBS + 0.05% BSA before incubating at room temperature for 30 minutes. Beads were then incubated on a magnetic strip for 5 minutes before supernatant was removed for future analysis. Mock depletions were performed as above only with unconjugated beads. Depleted samples were confirmed by ELISA for both efficiency and selectivity of depletion.

### QUANTIFICATION AND STATISTICAL ANALYSIS

Data and statistical analyses were performed using GraphPad Prism 10.2.2. Flow cytometry data was analyzed using FlowJo 10.10. One-way Anova, Kruskal-Wallis test followed by Dunn’s multiple comparisons test, unpaired two-tailed t-test with Welch’s correction for unequal variances, and Mann Witney two-tailed T-tests were used in the indicated figures. Statistical significance was defined as *p* < 0.05.

---

**Figure S1 (related to Fig. 1):**
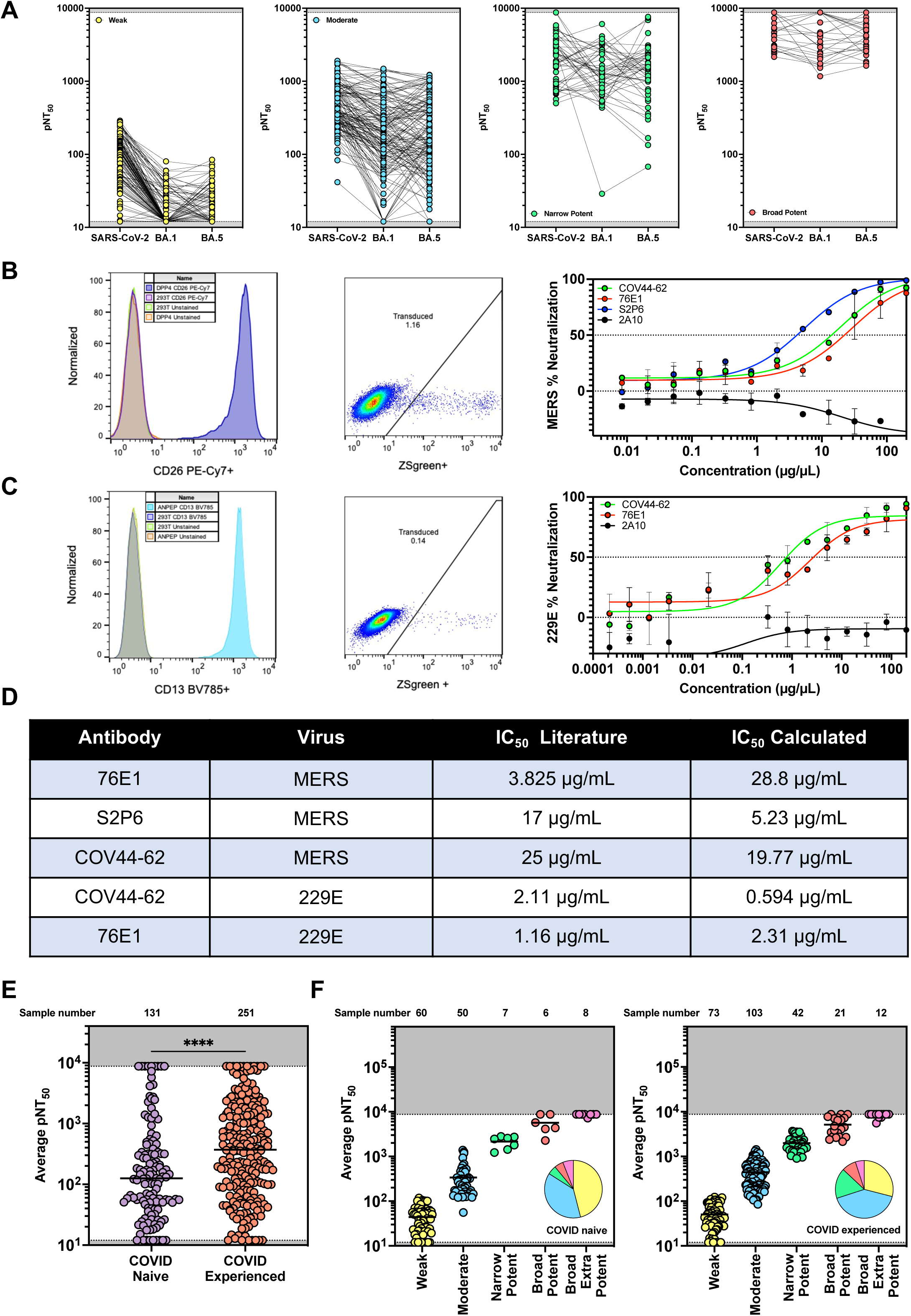
Individual neutralization values for donor cohort and development and validation of MERS and 229E neutralization assays. (A) Neutralization Assay results (average of n = 2 technical replicates) against SARS-CoV-2 WT and BA.1, BA.5 variants for a cohort of 382 donor samples. (B-C) Adaptation of our high throughput neutralization assay to evaluate MERS (B) and 229E (C) neutralization. We transduced 293T cells to express either hDPP4 (B) or hANPEP (C) and confirmed cell-surface expression by flow cytometry (Left). These cell lines were tested for transduction by MERS (B) or 229E (C) pseudoviruses, respectively (Middle). Finally, we performed neutralization assays with known neutralizing antibodies COV44-62, 76E1, and S2P6, and 2A10 as a negative control to evaluate these cell lines and pseudoviruses (Right). Data represented as mean +/- SD. (D) Table comparing the IC_50_ values for antibodies in the literature relative to IC_50_ values we calculated. (E) Average neutralizing activity of 382 serum samples from donors who were either COVID-naïve or previously infected. Because the data did not follow a normal distribution (Shapiro-Wilk p < 0.0001), differences between COVID-naïve and COVID-experienced groups were analyzed using the Mann-Whitney U test. ****= P<0.0001. (F) Samples from naïve donors (left) or COVID-experienced donors (right) were divided into five groups based on neutralization potency, as defined in Figure 1A. Pie charts represent the proportion of each group.

**Figure S2 (related to Fig. 2):**
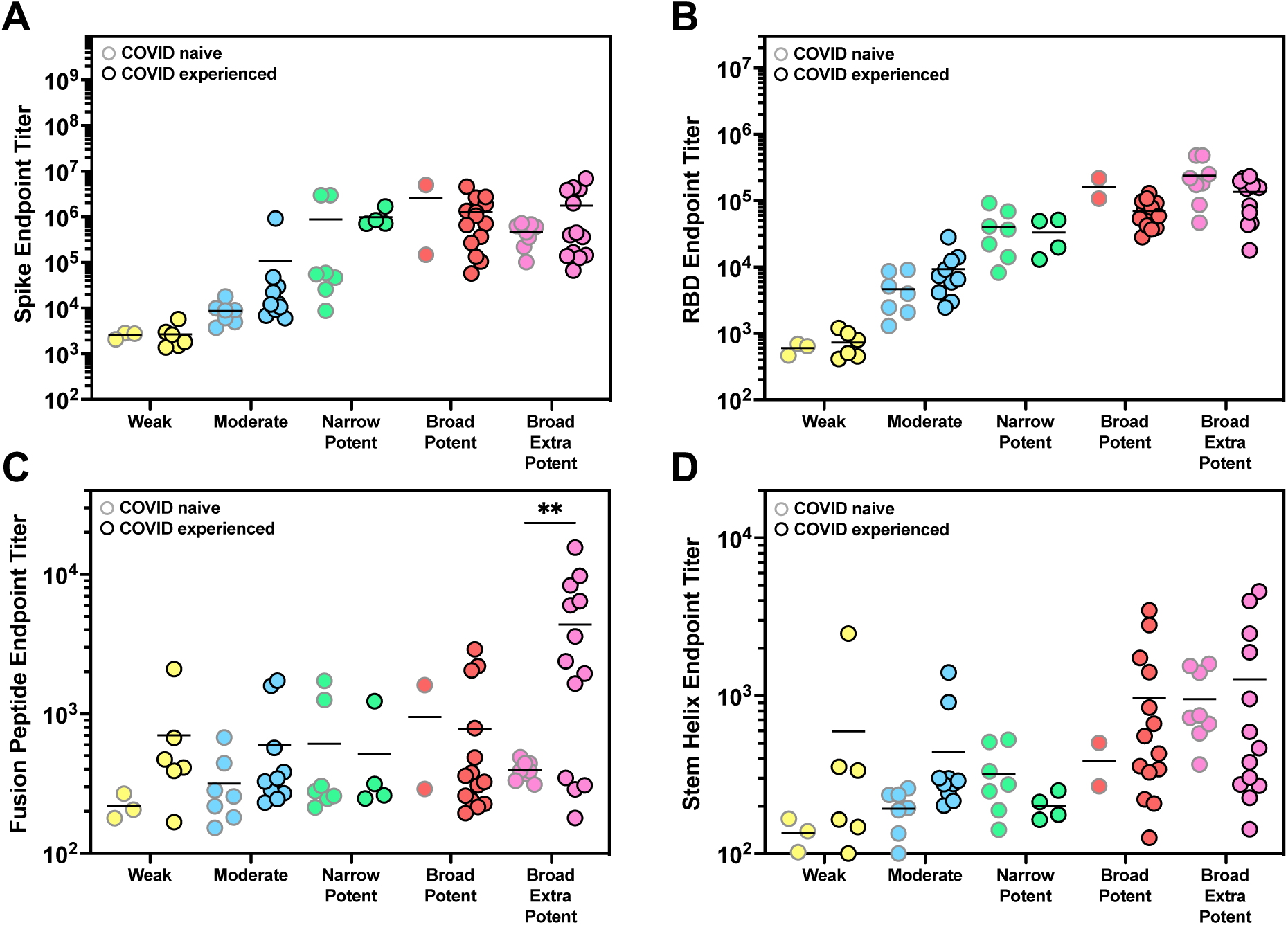
Broad extra potent samples from naïve donor have higher levels of FP antibodies. (A-D) Data from Figure 2B-E were further divided into two groups based on COVID history. Samples from COVID-naïve donors were outlined in gray, and those from COVID-experienced donors were outlined in black. Endpoint titers were compared between each COVID-naïve and COVID-experienced group using an unpaired two-tailed t-test with Welch’s correction for unequal variances. ** = P<0.0021.

**Figure S3 (related to Fig. 3):**
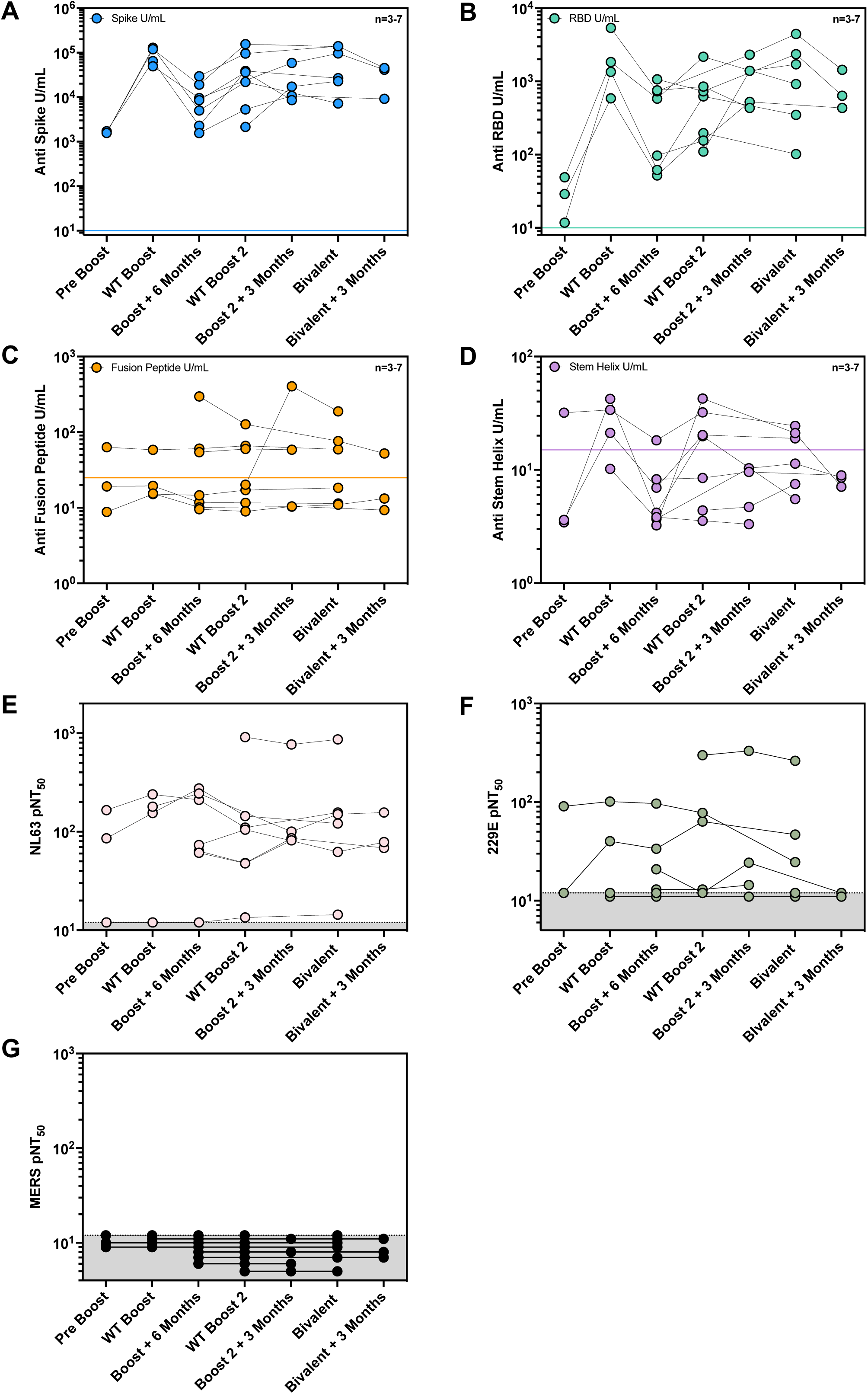
Repeated vaccination does not correlate with antigen specific antibodies, nor does it increase neutralization against alpha coronaviruses or MERS. (A-D) ELISA (average of n = 2 technical replicates) results for anti-SARS-CoV-2 Spike (A), anti-RBD (B), anti-Fusion Peptide (C), and anti-Stem Helix (D) antibodies. One U/mL is defined as the equivalent reactivity seen by 1 µg/mL RBDCOV182 antibody or 1 µg/mL S2P6 for SH or 1 µg/mL 76E1 for FP. Each line corresponds to a single donor over time. (E-G) Neutralizing titer (average of n = 2 technical replicates) against NL63 (E), 229E (F), or MERS (G) was assessed for longitudinal timepoints. Each line corresponds to a single donor over time.

**Figure S4 (related to Fig, 4):**
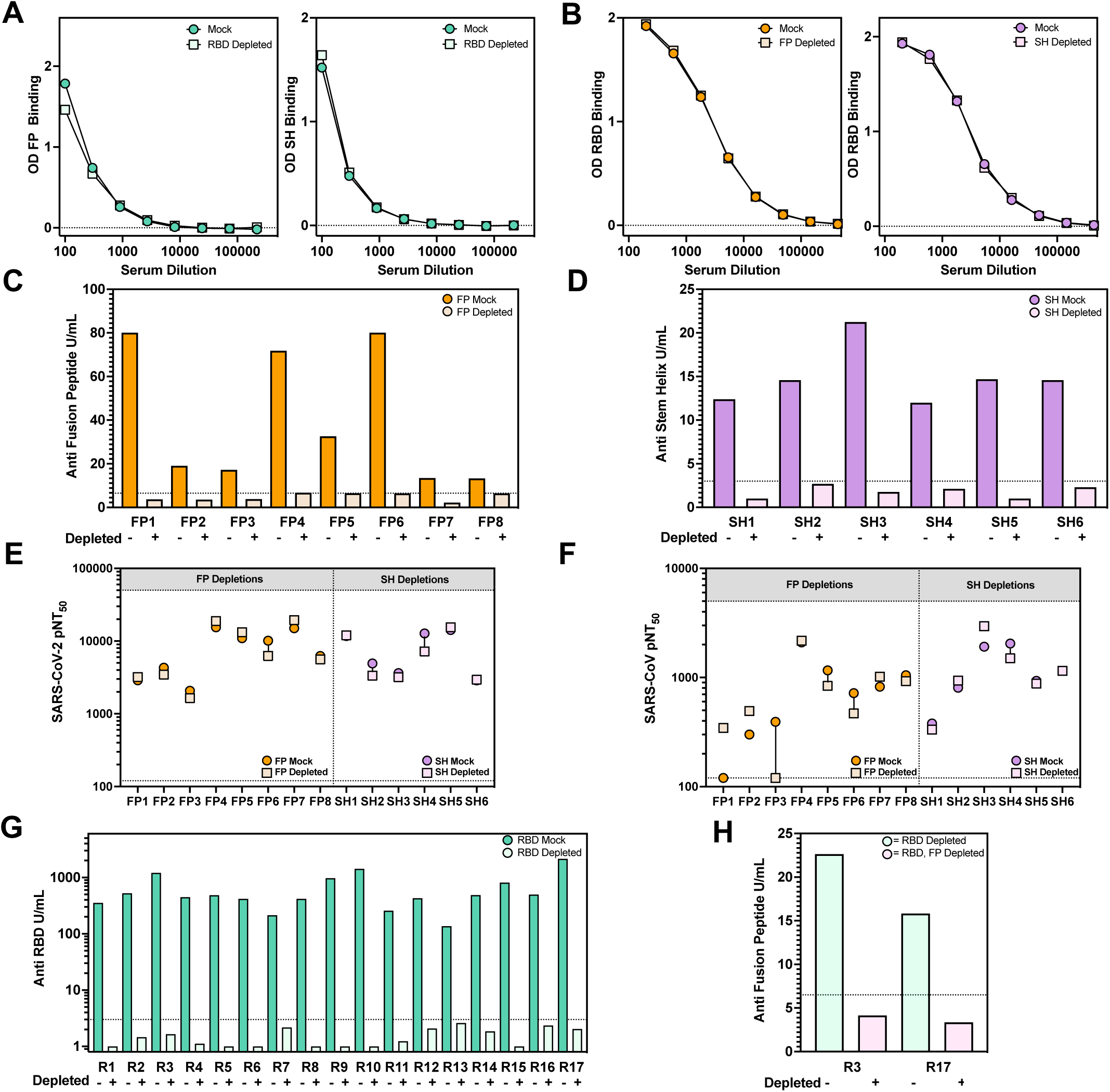
Antigen depletions effectively remove antigen specific antibodies. (A) Selectivity of RBD depletion. Serum samples were subject to Mock or RBD depletion and then tested by ELISA (average of n = 2 technical replicates) for binding to either the Fusion Peptide (left) or Stem Helix (right). Two representative samples are shown for each depletion for samples that had strong binding to either the FP or the SH. (B) Selectivity of Fusion Peptide or Stem Helix Depletion. Serum samples were subject to depletion against either the FP (left) or the SH (right) and then tested for binding to RBD by an ELISA (average of n = 2 technical duplicates). Two representative samples are shown for each depletion. (C-D) Depletion ELISA detailing the effect of FP (C) or SH (D) depletion on 6-8 broadly neutralizing samples. Samples with high FP or SH binding were chosen for depletion with those antigens. U/mL is calculated with 1 U/mL defined as the equivalent reactivity seen by 1 µg/mL, 76E1^43^ for FP and S2P6^41^ for SH. Cutoffs were defined by a negative control antibody (2A10). (E-F) Serum samples were subject to either fusion peptide or stem helix depletion to remove peptide specific antibodies and then tested for neutralizing activity (average of n = 2 technical replicates) to either SARS-CoV-2 (E) or SARS-CoV (F). Mock samples were combined with beads only for mock depletion. (G) Depletion ELISA (average of n = 2 technical replicates) detailing the effect of RBD depletion on 17 broadly neutralizing samples. Samples with high FP or SH binding were chosen for depletion with those antigens. U/mL is calculated with 1 U/mL defined as the equivalent reactivity seen by 1 µg/mL RBDCOV182 (isolated in house) antibody for RBD. Cutoffs were defined by a negative control antibody (2A10). (H) RBD Depleted donors with remaining neutralizing activity were subject to fusion peptide depletion and an ELISA (average of n = 2 technical replicates) was done to confirmed depletion. Cutoffs were defined by a negative control antibody (2A10).

**Figure S5 (related to Fig. 5):**
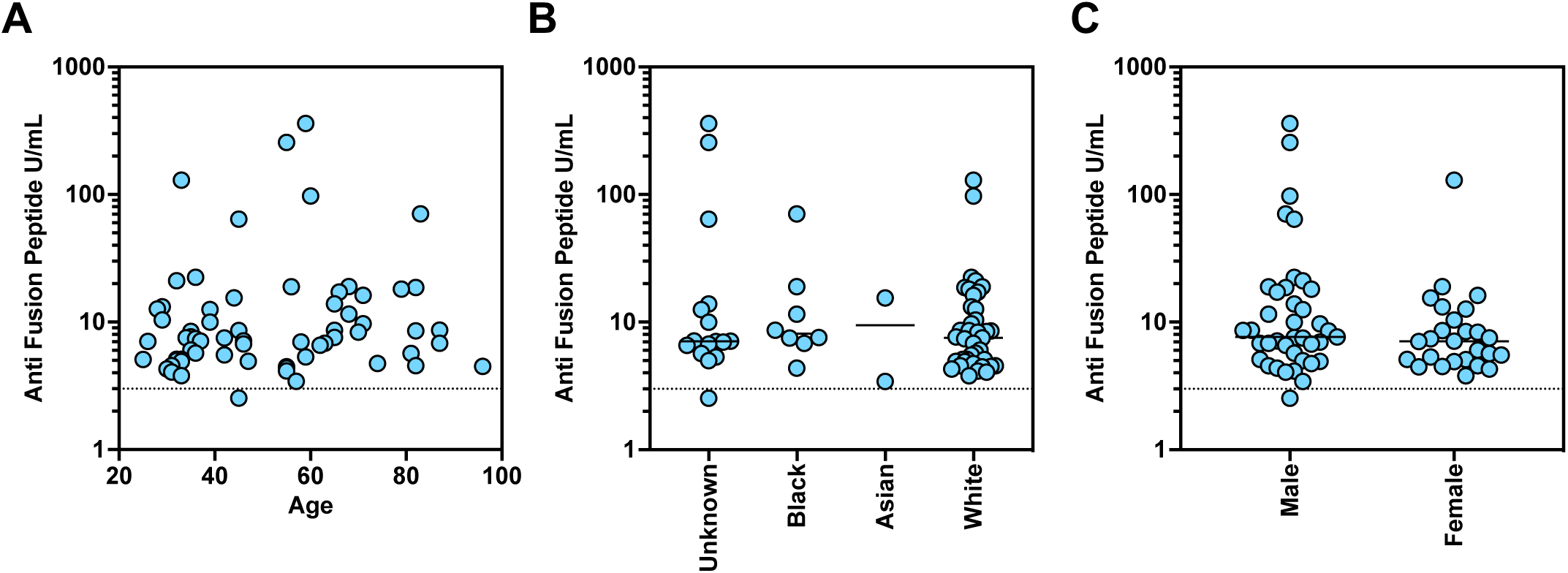
Age, race, and biological sex did not correlate with FP antibody development. (A-C) Fusion peptide U/mL ELISA (average of n = 2 technical replicates) for COVID-experienced donors separated by age (A), race (B), biological sex (C). Age, race and biological sex were not correlated with FP-specific antibodies. For A-C U/mL is defined as the reactivity seen by 1 µg/mL of 76E1^43^ antibody. Cutoffs were determined by binding of a negative control antibody (2A10).

**Table S1 (related to Fig 1A):** Cohort Descriptions for Figure 1A. Descriptions of the various groups for Figure 1A and Supplemental Figure 1A with age and sex noted.

| Cohort | Statistics |
| --- | --- |
| **Total (n)** | 382 |
| **Broad Extra Potent** | 16 |
| Age (y) (median, range) | 70 (57-77) |
| Biological Sex (% Female) | 7/16 (44%) |
| COVID History (% Experienced) | 11/16 (69%) |
| **Broad Potent** | 31 |
| Age (y) (median, range) | 73 (57-93) |
| Biological Sex (% Female) | 20/31 (65%) |
| COVID History (% Experienced) | 21/31 (68%) |
| **Narrow Potent** | 49 |
| Age (y) (median, range) | 77 (40-97) |
| Biological Sex (% Female) | 32/49 (65%) |
| COVID History (% Experienced) | 42/49 (86%) |
| **Moderate** | 153 |
| Age (y) (median, range) | 76 (33-100) |
| Biological Sex (% Female) | 62/153 (41%) |
| COVID History (% Experienced) | 103/153 (67%) |
| **Weak** | 133 |
| Age (y) (median, range) | 73 (49-100) |
| Biological Sex (% Female) | 23/133 (17%) |
| COVID History (% Experienced) | 73/133 (55%) |

**Table S2 (related to Fig 1A):** Collection TimePoints of Samples Analyzed in Figure 1A. Number of samples collected at each time point. All donors received one vaccination followed by at least one booster dose. Sera were collected either before the booster (pre-boost) or at 2 weeks, 3 months, 6 months, or 12 months after vaccination.

| Timing of Sampling | Total | Weak<br>(n=133) | Moderate<br>(n=153) | Narrow<br>Potent<br>(n=49) | Broad<br>Potent<br>(n=27) | Broad<br>Extra<br>Potent<br>(n=20) |
| --- | --- | --- | --- | --- | --- | --- |
| Pre-boost | 16 | 11/16 (69%) | 5/16 (31%) | 0/16 (0%) | 0/16 (0%) | 0/16 (0%) |
| 2 Weeks After First Boost | 39 | 3/39 (8%) | 19/39 (49%) | 9/39 (23%) | 5/39 (13%) | 3/39 (8%) |
| 3 Months After First Boost | 27 | 16/27 (59%) | 10/27 (37%) | 0/27 (0%) | 1/27 (4%) | 0/27 (0%) |
| 6 Months After First Boost | 99 | 47/99 (47%) | 40/99 (40%) | 7/99 (7%) | 3/99 (3%) | 2/99 (2%) |
| 12 Months After First Boost | 57 | 40/57 (70%) | 16/57 (28%) | 0/57 (0%) | 1/57 (2%) | 0/57 (0%) |
| 2 Weeks After Second Boost | 70 | 1/70 (1%) | 31/70 (44%) | 18/70 (26%) | 16/70 (23%) | 4/70 (6%) |
| 3 Months After Second Boost | 67 | 15/67 (22%) | 32/67 (48%) | 15/67 (22%) | 1/67 (1%) | 4/67 (6%) |
| 2 Weeks After Third Boost | 5 | 0/5 (0%) | 0/5 (0%) | 0/5 (0%) | 0/5 (0%) | 5/5 (100%) |
| 3 Months After Third Boost | 2 | 0/2 (0%) | 0/2 (0%) | 0/2 (0%) | 0/2 (0%) | 2/2 (100%) |

**Table S3 (related to Fig S1E):**
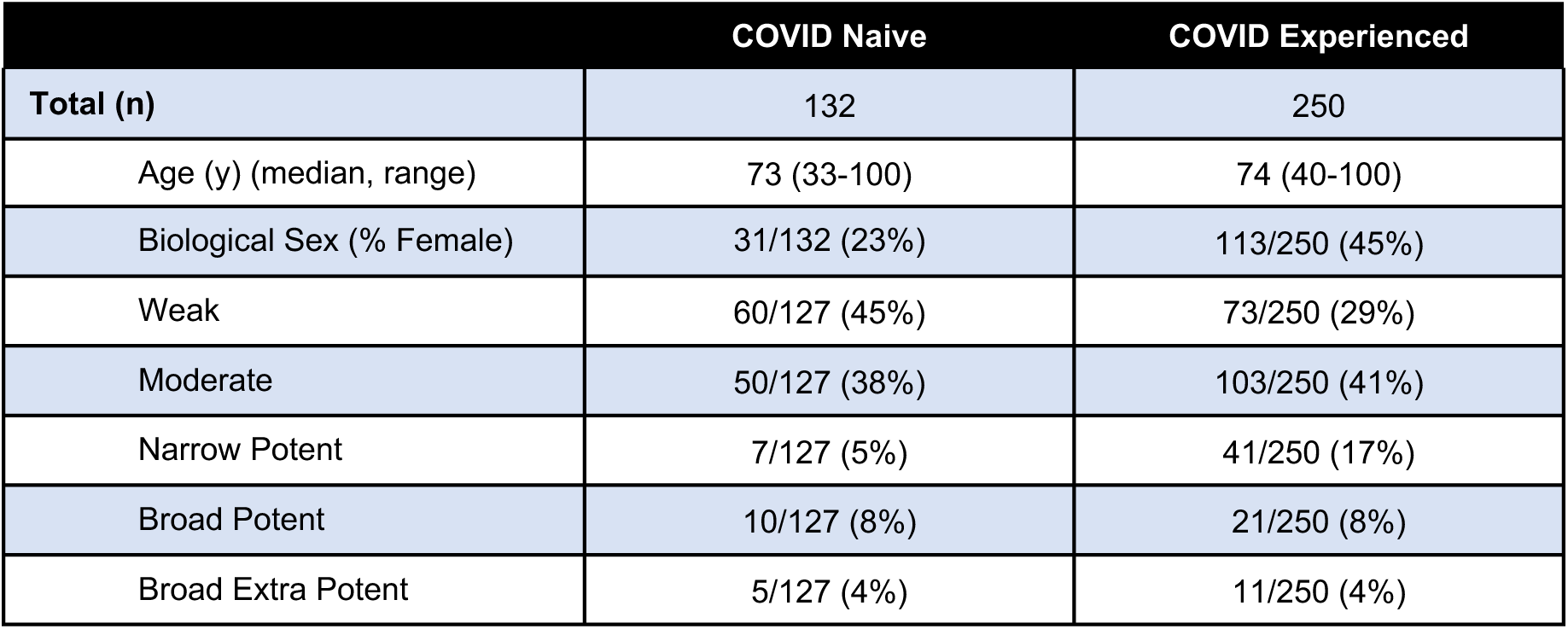
Comparison of Samples from COVID-Naïve or Experienced Donors Analyzed in Supplemental Figure 1E. Descriptions of samples from donors who were either COVID-naïve or previously infected with age and sex noted. Proportion of sample numbers in each groups were also analyzed.

**Table S4 (related to Fig 3, 4):** Demographics of longitudinal donor cohort. Table describing individual timepoints from 8 of the broadest neutralizing donors spanning 3-5 timepoints including those 2-4 weeks after vaccination as well as timepoints that were distant (Post) from the nearest vaccination (3-6 months post) from a cohort of Nursing Home Samples.

| Donor | Age | Biological Sex | Vaccination Status | Infection History |
| --- | --- | --- | --- | --- |
| 1 | 81-85 | Male | 4XWT, 1XBivalent | Infection Prior to 1<sup>st</sup> Vax |
| 2 | 66-70 | Female | 4XWT, 1XBivalent | Naive |
| 3 | 55-60 | Male | 4XWT, 1XBivalent | Infection Prior to 1<sup>st</sup> Vax |
| 4 | 61-65 | Male | 4XWT, 1XBivalent | Naive |
| 5 | 61-65 | Female | 4XWT, 1XBivalent | Infection After 4<sup>th</sup> Vax |
| 6 | 66-70 | Female | 4XWT, 1XBivalent | Infection Prior to 1<sup>st</sup> Vax |
| 7 | 86-90 | Female | 4XWT, 1XBivalent | Naive |
| 8 | 76-70 | Male | 4XWT, 1XBivalent | Infection Prior to 1<sup>st</sup> Vax<br>Infection after 4<sup>th</sup> Vax |

**Table S5 (related to Fig 5):** Cohort Descriptions for Figure 5. Descriptions of the various cohorts for Figure 5 and Supplemental Figure 5 with age and sex noted.

| Cohort | Statistics |
| --- | --- |
| **Total (n)** | 255 |
| **Demographics All Cohorts** |  |
| Age (y) (median, range) | 69 (25-96) |
| Biological Sex (% Female) | 93/255 (36%) |
| **COVID+ Pre-Vaccination Total (n)** | 74 |
| Age (y) (median, range) | 63 (25-96) |
| Biological Sex (% Female) | 33/94 (35.1%) |
| **COVID Naïve Pre-Vaccine Total (n)** | 14 |
| Age (y) (median, range) | 70 (60-88) |
| Biological Sex (% Female) | 3/14 (21%) |
| **COVID Experienced Pre-Vaccine Total (n)** | 71 |
| Age (y) (median, range) | 55 (25-96) |
| Biological Sex (% Female) | 29/71 (41%) |
| **COVID Naïve Post-Vaccine (5C) Total (n)** | 72 |
| Age (y) (median, range) | 73 (49-89) |
| Biological Sex (% Female) | 35/72 (49%) |
| **COVID Experienced Post-Vaccine (5C) Total (n)** | 52 |
| Age (y) (median, range) | 70 (37-88) |
| Biological Sex (% Female) | 13/52 (25%) |
| **COVID Naïve Longitudinal (5D) Total (n)** | 30 |
| Age (y) (median, range) | 73 (49-88) |
| Biological Sex (% Female) | 10/30 (33%) |
| **Longitudinal BT Cohort (5E) Total (n)** | 7 |
| Age (y) (median, range) | 73 (65-88) |
| Biological Sex (% Female) | 2/7 (29%) |

## Notes

### Author Declarations

Use of human samples was approved by Partners Institutional Review Board (protocol 2020P002274).

### Summary of Updates

This version of the manuscript has been revised to respond to reviews.

